# A three-gene expression score for predicting clinical benefit to anti-PD-1 blockade in advanced renal cell carcinoma

**DOI:** 10.1101/2023.12.03.23299342

**Authors:** Yoel Z. Betancor, Miriam Ferreiro-Pantín, Urbano Anido-Herranz, Mar Fuentes-Losada, Luis León-Mateos, Silvia Margarita García-Acuña, Vanessa Vaamonde-Rodríguez, Beatriz García-Pinel, Víctor Cebey-López, Rosa Villaverde-Viaño, Martín Kotrulev, Natalia Fernández-Díaz, Iria Gomez-Tourino, Carlos Fernández-Baltar, Jorge García-González, Jose M. C. Tubio, Rafael López-López, Juan Ruiz-Bañobre

**Author notes:** **Corresponding author:** Dr. Juan Ruiz-Bañobre; Medical Oncology Department, University Clinical Hospital of Santiago de Compostela, Travesía da Choupana S/N, 15706 Santiago de Compostela, Spain. Co-senior authors (Rafael López-López, Juan Ruiz-Bañobre). These authors contributed equally to this work (Miriam Ferreiro-Pantín, Urbano Anido-Herranz).

## Abstract

**Background:** In the advanced renal cell carcinoma (RCC) scenario there are no consistent biomarkers to predict the clinical benefit patients derived from immune checkpoint blockade. We conducted a retrospective study in order to develop and to validate a gene expression score for predicting clinical benefit to nivolumab in the advanced clear cell RCC setting and to characterize its underlying clinical, molecular, and immune features.

**Methods:** This is a *post hoc* pooled analysis of 311 patients with available clinical, molecular, and immune tumor data from the CheckMate-009, -010 and -025 trials. Efficacy endpoints were overall survival (OS), disease control rate, and overall response rate. Survival estimates were calculated by the Kaplan-Meier method, and groups were compared with the log-rank test. The Cox proportional hazards regression model was used to evaluate factors independently associated with OS. Factors associated with disease control and response were tested with logistic regression. Biomarker-treatment interaction was evaluated with the likelihood ratio test (LRT).

**Results:** First, a three-gene expression score (3GES) with prognostic value for OS integrating HMGA1, NUP62, and ARHGAP42 transcripts was developed in a cohort of patients treated with nivolumab. Favorable 3GES risk category was significantly associated with a better OS in univariable (HR = 0.32, 95% CI 0.21 - 0.48, *P*<0.001) and multivariable (HR = 0.36, 95% CI 0.24 - 0.56, *P*<0.001) analyses. Consistent and significant correlation was found with disease control (*P*=0.066) and response (*P*=0.002). The 3GES prognostic value was validated in the TCGA-KIRC cohort (HR=0.35, *P*<0.001). Next, the predictive value for nivolumab was confirmed in a set of patients from the CheckMate-025 trial (LRT *P*<0.001).

**Conclusions:** In accRCC, 3GES is not only an independent prognostic factor for OS but also a positive predictive biomarker for nivolumab. The predictive value deserves further validation in other retrospective and large-scale, randomized prospective studies.

## Introduction

The emergence of immunotherapy, particularly programmed cell death-1 (PD-1) antibodies, has revolutionized the management of patients diagnosed with clear cell renal cell carcinoma (ccRCC) over the last decade. Since the approval of nivolumab (an anti-PD-1 antibody) in November 2015 for the treatment of patients with advanced ccRCC (accRCC) who received previous antiangiogenic therapy, new immunotherapy-based combination regimens have been approved for previously untreated patients^1^. Today, in the first-line setting, there are several immune checkpoint blockade (ICB)-based alternatives that have been approved by the main regulatory agencies after demonstrating significant overall survival improvements in randomized, phase 3 clinical trials: pembrolizumab (an anti-PD-1 antibody) plus either axitinib or lenvatinib (both tyrosine kinase inhibitors)^2,3^, nivolumab plus cabozantinib (a tyrosine kinase inhibitor)^4^, and nivolumab plus ipilimumab (an anti-CTLA-4 antibody) (this combination only for those patients with intermediate or poor-risk aRCC)^5^. In the second and subsequent lines of therapy, there are available different single-agent options beyond nivolumab such as tyrosine kinase- and mTOR-inhibitors^5^. Furthermore, very recently, pembrolizumab has been approved for the treatment of adults with ccRCC at increased risk of recurrence following nephrectomy or following nephrectomy and resection of metastatic lesions^6^.

In parallel to the growth of the immunotherapeutic armamentarium, to identify ccrRCC patients most likely to benefit from ICB has become a priority. Over the last years, a plethora of studies have evaluated the role of different prognostic and/or predictive biomarkers for ICB in accRCC. Numerous translational research initiatives have explored the role of different molecular markers such as PD-L1^1,7,8^, tumor mutational burden (TMB)^9–12^, PBRM1 loss-of-function mutations^13^, alterations in DNA damage response and repair genes^13^, gene expression signatures^13^, and T-cell receptor clonality in the tumor microenvironment^14^. Other host-related biomarkers such as obesity^13,15^, presence of pancreatic metastases^16^, the International Metastatic RCC Database Consortium (IMDC) Risk Score^7^, or the gut microbiome have also been evaluated. Nevertheless, to date, IMDC Risk Score is the only biomarker used in the clinical practice as a selection criterion to treat patients with the combination regimen of nivolumab plus ipilimumab^7^.

Taking this into consideration, herein we conducted a retrospective study in order to develop and to validate a gene expression score for predicting clinical benefit to the anti-PD-1 antibody nivolumab in the context of patients diagnosed with accRCC enrolled in the CheckMate-009, -010 and -025 clinical trials. Additionally, we explored the correlation of our three-gene expression score (3GES) with different clinical, molecular, and immune tumor characteristics.

## Patients and Methods

### Study design and patient population

The design and primary outcomes of the CheckMate-009, -010 and -025 trials were described in previous reports^1,17,18^. Briefly, CheckMate-009^17^ was an open-label, parallel, four-group, phase 1 trial that investigated the pharmacodynamic immunomodulatory activity, efficacy, and safety of nivolumab in patients with previously treated accRCC; CheckMate-010 was a blinded, randomized, multicenter phase 2 trial that evaluated the dose-response relationship, efficacy, and safety of nivolumab in patients with previously treated acRCC^18^; and CheckMate-025 was a two-arm, randomized, open-label, phase 3 study that compared nivolumab with everolimus in patients with previously treated accRCC^1^. This is a *post hoc* pooled analysis of 311 patients with available clinical, molecular, and immune tumor data from the CheckMate-009, -010 and -025 trials (16, 45, and 250 patients, respectively)^1,17,18^. For the purpose of our analyses, our efficacy endpoints were overall survival (OS), disease control rate (DCR), and overall response rate (ORR). Tumor responses were assessed according to Response Evaluation Criteria in Solid Tumors guidelines version 1.1. Additionally, the TCGA Clear Cell Renal Cell Carcinoma (TCGA-KIRC) cohort was used as an external validation set.

All clinical and molecular tumor data (generated from pre-treatment tumor samples) used for this retrospective study have been made freely available through Supplementary Material by Braun et al^9^. Briefly, RNA-seq data from the CheckMate-010 and -025 cohorts were aligned using STAR^19^, quantified using RSEM^20^, and evaluated for quality using RNA-seQC2^21^. Samples were excluded if they had an interquartile range of log_2_[transcript per million (TPM) + 1] < 0.5 (indicating low dynamic range), had less than 15,000 genes detected (indicating low library complexity), had an End 2 Sense Rate<0.90, or End 1 Sense Rate>0.10 (as defined by RNA-seqQC2, indicating strand bias). For samples where RNA-seq was performed in duplicates, the run with a higher interquartile range of log_2_(TPM + 1), used as a surrogate for better quality data, was used. For the CheckMate-009 cohort, the previously published TPM matrix was used^22^. Genes that were not expressed in any of the samples (in each cohort independently) then upper quartile-normalized the TPMs to an upper quartile of 1000, and log_2_-transformed them were filtered. Since the sequencing had been performed in 4 separate batches, principal component analysis (PCA) was used to evaluate for batch effects and 4 batches were observed. These 4 batches were corrected by using Combat^23^. Subsequently, a PCA was performed on the ComBat-corrected expression matrix to confirm that batch effects had been adequately corrected for. Moreover, a constant that was equal to the first integer above the minimum negative expression value obtained post-ComBat (constant of +21) to eliminate negative gene expression values that were a by-product of ComBat correction. The ComBat-corrected expression matrix was used for all downstream analyses. RNA-seq data from the TCGA-KIRC were downloaded from the UCSC Xena Browser (dataset identification: TCGA.KIRC.sampleMap/HiSeqV2) as log_2_(normalized_count+1) and transformed to TPM values. The expression levels of all the genes were independently dichotomized for each cohort into high and low using the maximally selected rank statistic (maxstat.test() function from *maxstat* R package. Computational immune cell deconvolution was carried out with EPIC version 1.1.7 R package^24^.

Statements confirming compliance with ethical regulations, the committees that approved the protocol of CheckMate studies, and confirmation of informed consent from all study participants are included in the previous publications describing these trials (NCT01358721, NCT01354431, and NCT01668784)^1,17,18^.

### Statistical analysis

Survival estimates were calculated by the Kaplan-Meier method, and groups were compared with the log-rank test. The Cox proportional hazards regression model was used to evaluate factors independently associated with OS. Baseline clinicopathological variables included in the multivariable analysis were selected according to statistical significance in univariable analysis (cutoff, *P* < 0.05) (**Supplementary Table 1**). The proportional hazard assumption was verified with the Schoenfeld residual method. Factors associated with disease control (DC) and response were tested with logistic regression in univariable analyses. Variables included in the final multivariable model were selected according to their statistical significance in univariable analysis (cutoff, *P* < 0.05). Biomarker-treatment interaction was evaluated with the likelihood ratio test. Comparisons between patient and disease characteristics were carried out using Chi-squared or Fisher exact tests. Comparisons between estimated cell fractions and immune exhaustion markers expression levels were carried out using Wilcoxon rank-sum test. All *P* values were 2-sided, and those less than 0.05 were considered statistically significant. The Bonferroni and the Benjamini–Hochberg (B-H) procedures were used to control the family-wise error rate (FWER) and the false discovery rate (FDR), respectively, in case of multiple comparisons. All statistical analyses were performed using R version 4.2.2 (Vienna, Austria).

## Results

### Development and validation of a novel three-gene expression score with prognostic significance

First, to identify genes with prognostic significance among ccRCC patients treated with immunotherapy, we evaluated the association of 43893 transcripts with OS by using univariable Cox Proportional Hazard models in a pooled cohort of 181 patients treated with nivolumab from the CheckMate-009, -010 and -025 trials. Seventeen out of the 43893 transcripts evaluated were significantly associated with OS (Bonferroni FWER-adjusted *P* < 0.05) (**Figure 1A** and **Supplementary Table 2**). Next, to further optimize the selection of genes, we filtered out those transcripts that exhibited no correlation with either disease control or response. Seven out of 17 genes showed a statistically significant association with either disease control, response, or both (**Figure 1A** and **Supplementary Table 3**). Importantly, the direction of this association for each gene was clinically consistent with that previously identified with OS. Lastly, we used a multivariable Cox Proportional Hazard model to obtain a final panel of 3 independent prognostic genes (**Figure 1A** and **Supplementary Table 4**). A high expression of HMGA1 and NUP62 was associated with a worse OS (HR = 1.60, 95% CI 1.05 - 2.46, *P* = 0.031; and HR = 1.74, 95% CI 1.17 - 2.60, *P* = 0.007, respectively) (**Supplementary Table 4**). On the contrary, a low expression of ARHGAP42 was associated with a worse OS (HR = = 1.74, 95% CI 1.15 - 2.63, *P* = 0.008) (**Supplementary Table 4**).

**Figure 1.**
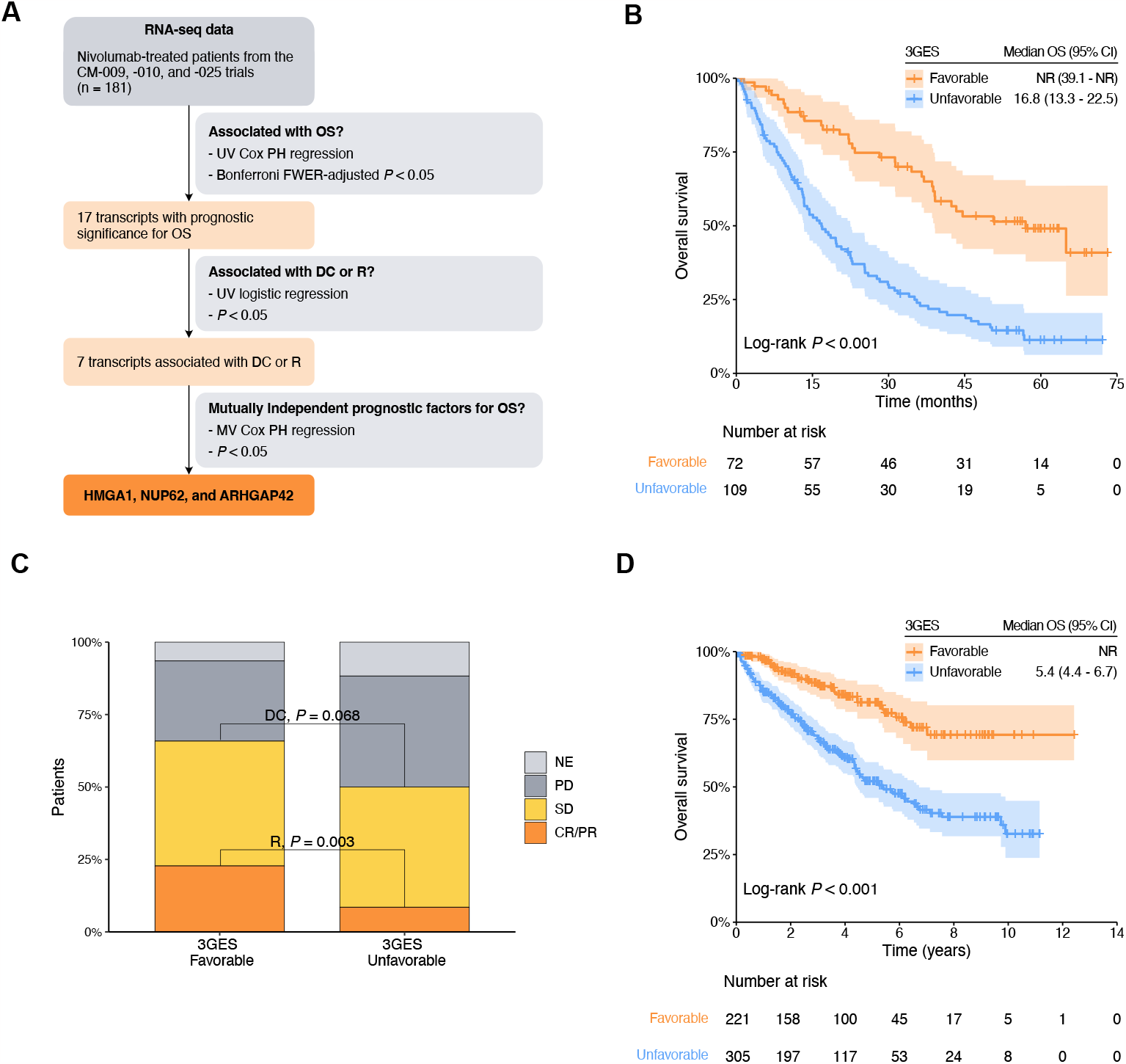
**(A)** Flow diagram of the selection process of final transcripts integrated into the three-gene expression score (3GES). **(B)** Kaplan–Meier overall survival estimates of nivolumab treated patients from the pooled cohort of the CheckMate-009, -010, and -025 trials according to the 3GES. **(C)** Nivolumab response distribution by 3GES. **(D)** Kaplan–Meier overall survival estimates of patients from the TCGA-KIRC cohort. Abbreviations: 3GES, 3-gene expression score; CI, confidence interval; CM, CheckMate; CR, complete response; DC, disease control; FWER, family-wise error rate; NE, not evaluable; NR, not reached; OS, overall survival; PD, progressive disease; PH, proportional hazards; PR, partial response; R, response; SD, stable; UV, univariable.

Considering the amount of these 3 adverse prognostic genes (high HMGA1 = 1 point, high NUP62 = 1 point, and low ARHGAP42 = 1 point), we developed a 3GES to segregate patients into two risk categories. Patients without any adverse prognostic gene (0 points) were classified in the favorable-risk category [40%, n = 72 ; median OS = not reached (NR) (95% CI, 39.1 - NR)], and patients with one or more adverse prognostic genes (1 to 3 points) were classified in the unfavorable-risk category [60%, n = 109 ; median OS = 16.8 months (95% CI, 13.3 - 22.5)]. The Kaplan–Meier curves depicting these two risk categories are presented in **Figure 1B**. Favorable 3GES risk category was significantly associated with a better OS in univariable (HR = 0.32, 95% CI 0.21 - 0.48, *P* < 0.001) and multivariable (HR = 0.36, 95% CI 0.24 - 0.56, *P* < 0.001) analyses (**Table 1**). DCR and ORR were higher among favorable 3GES risk category patients (DCR for favorable *vs* unfavorable 3GES risk category patients: 65% *vs* 51%, *P* = 0.068; ORR for favorable *vs* unfavorable 3GES risk category patients: 33% *vs* 14%, *P* = 0.003; **Figure 1C**). Moreover, favorable 3GES risk category patients presented a higher probability of disease control and response in univariable [DC: odds ratio (OR) = 1.78, 95% CI 0.96 - 3.29, *P* = 0.066; response: OR = 3.13, 95% CI 1.51 - 6.54, *P* = 0.002] and multivariable (DC: OR = 1.85, 95% CI 0.93 - 3.70, *P* = 0.081; response: OR = 3.12, 95% CI 1.39 - 6.98, *P* = 0.006) analyses (**Table 1**).

**Table 1.**
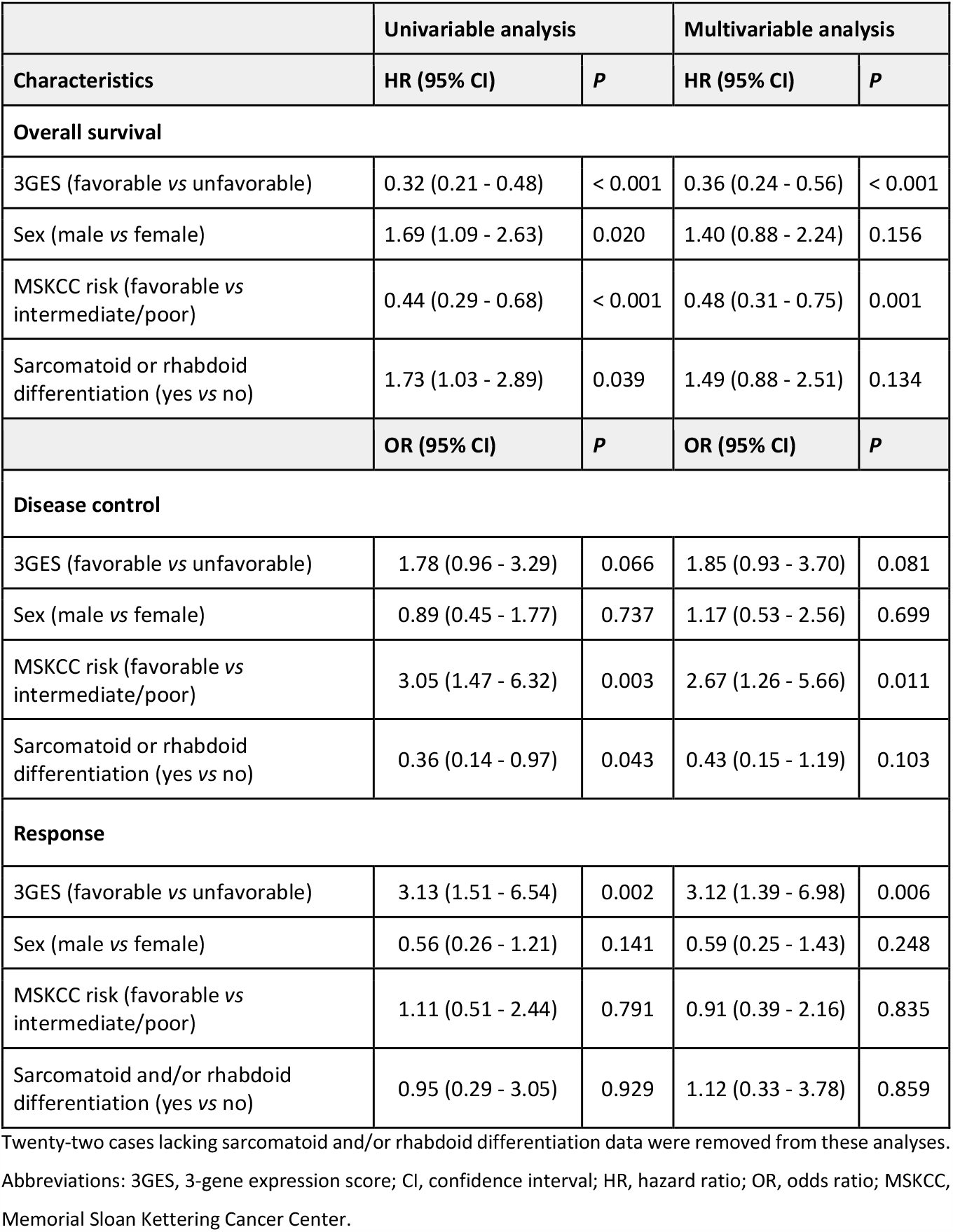
Univariable and multivariable Cox regression analyses for overall survival and logistic regression analyses for disease control and response among nivolumab treated patients included in the pooled cohort of the CheckMate-009, -010, and -025 trials..

Once the 3GES was developed, we went to validate its prognostic significance in an independent dataset, the TCGA-KIRC cohort. The Kaplan–Meier curves depicting these two risk categories are presented in **Figure 1D**. Consistent with our previous findings, favorable 3GES risk category was significantly associated with a better OS in univariable (HR = 0.35, 95% CI 0.24 - 0.50, *P* < 0.001) and multivariable (HR = 0.33, 95% CI 0.22 - 0.50, *P* < 0.001) analyses (**Table 2**).

**Table 2.** Univariable and multivariable Cox regression analyses for overall survival among patients from the TCGA-KIRC cohort.

|  | Univariable analysis |  | Multivariable analysis |  |
| --- | --- | --- | --- | --- |
| Characteristics | HR (95% CI) | *P* | HR (95% CI) | *P* |
| 3GES (favorable vs unfavorable) | 0.35 (0.24 - 0.50) | < 0.001 | 0.33 (0.22 - 0.50) | < 0.001 |
| AJCC stage at diagnosis (III-IV vs I-II) | 3.95 (2.87 - 5.45) | < 0.001 | 3.73 (2.57 - 5.42) | < 0.001 |
| Hemoglobin (low vs normal/high) | 2.15 (1.52 - 3.06) | < 0.001 | 1.36 (0.93 - 2.00) | 0.116 |
| Calcium (high vs normal/low) | 4.36 (2.20, 8.62) | < 0.001 | 2.28 (1.14 - 4.55) | 0.020 |

Three cases lacking AJCC stage at diagnosis data, 80 cases lacking hemoglobin data, and 164 cases lacking calcium data were removed from these analyses (number of cases included, n = 355).
Abbreviations: 3GES, 3-gene expression score; CI, confidence interval; HR, hazard ratio.

### Evaluation of the predictive value of the novel three-gene expression score

To further explore the predictive value of our 3GES when patients are treated with immunotherapy, we specifically interrogated those patients with available RNA-sequencing and clinical data from the CheckMate-025 study (n = 250). Among favorable 3GES risk category patients, nivolumab monotherapy significantly improved the OS compared to everolimus (nivolumab arm, median OS = NR, 95% CI 38.8 - NR *vs* everolimus arm, mOS = 32.8, 95% CI 24.7 - 43.4), with a reduction of death risk of 54% (HR = 0.46, 95% CI 0.27 - 0.79, *P* = 0.003) (**Figure 2A**). Conversely, among unfavorable 3GES risk category patients there was not significant differences in terms of OS based on the allocated treatment arm (nivolumab arm, median OS = 17.6, 95% CI 13.3 - 26.0 *vs* everolimus arm, mOS = 15.2, 95% CI 11.4 - 19.7; HR = 0.79, 95% CI 0.56 - 1.12, *P* = 0.19) (**Figure 2B**). Importantly, the 3GES-treatment interaction was statistically significant whether unadjusted (LRT *P* < 0.001, concordance = 0.64) or after adjustment for previously confirmed independent prognostic factors (sex, MSKCC risk group, and presence of sarcomatoid and/or rhabdoid histological differentiation) (LRT *P* < 0.001, concordance = 0.68).

**Figure 2.**
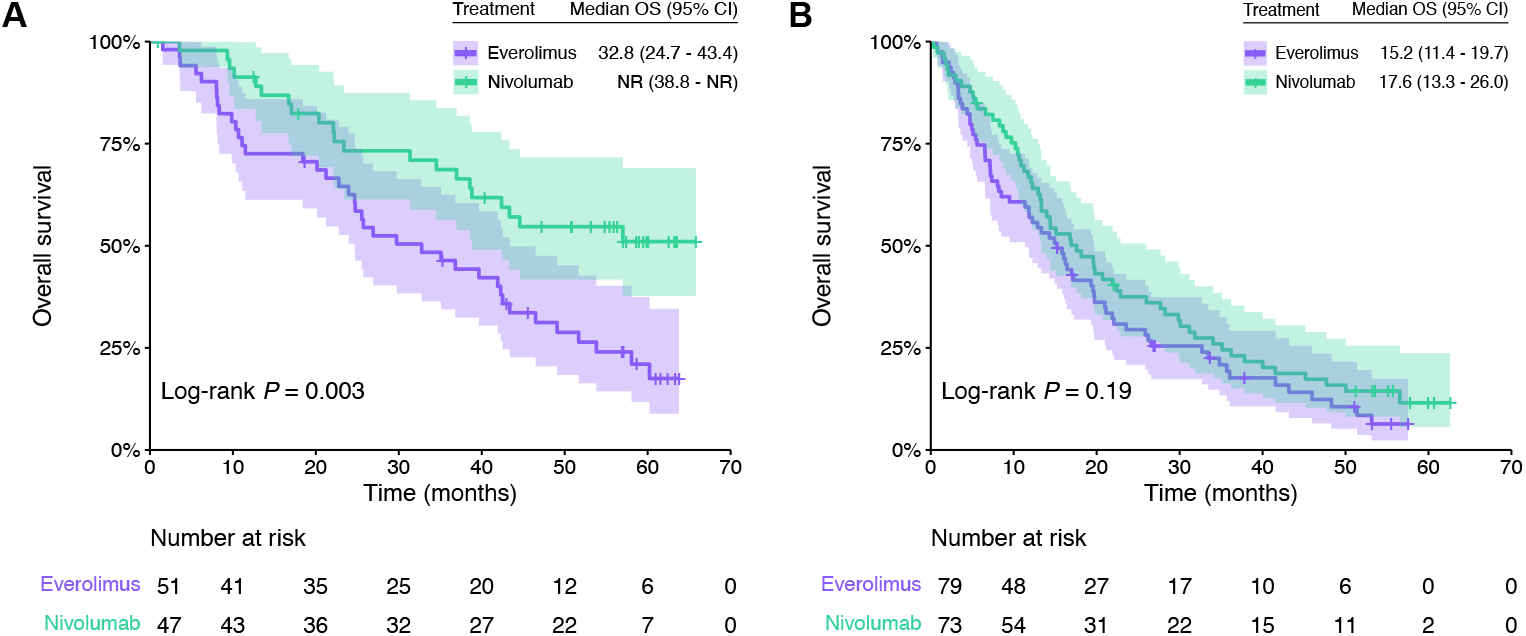
Kaplan–Meier overall survival estimates of patients from the CheckMate-025 trial according to the treatment arm (nivolumab *vs* everolimus) in **(A)** favorable and **(B)** unfavorable 3GES risk groups. Abbreviations: CI, confidence interval; OS, overall survival; NR, not reached.

### Clinical, molecular, and immune correlates of the three-gene expression score

To fully characterize our 3GES, we evaluated its correlation with available baseline patient and disease characteristics among the 311 subjects included in the CheckMate-009, -010 and -025 trial pooled cohort (**Supplementary Table 5**). Forty percent (n = 123) of patients had a favorable 3GES risk, while 60% (n = 188) had an unfavorable 3GES risk; 5% (n = 16) of patients come from CheckMate-009, 15% (n = 45) from CheckMate-010, and 80% (n = 250) from CheckMate-025. The distribution of different patient and disease characteristics according to the 3GES is shown in **Supplementary Table 5**. To note, in the favorable 3GES risk group there was a higher proportion of patients with favorable MSKCC risk group (*P* = 0.039), absence of sarcomatoid and/or rhabdoid histological differentiation (*P* = 0.042), and a lower copy number alteration burden [as measured by the weighted genome integrity index (wGII)] (*P* = 0.035). Importantly, there were no statistically significant differences in the cohort of origin (CheckMate-025 trial *vs* CheckMate-009 and -010 trials) among favorable and unfavorable 3GES risk cases.

Next, we evaluated whether any individual mutation or copy number alteration were associated with the 3GES. Interestingly, the tumors of patients from the favorable 3GES risk group presented a significant enrichment in *PBRM1* loss-of-function mutations (B-H FDR-adjusted *P* = 0.010) and the amplification 8Q24.3 (B-H FDR-adjusted *P* = 0.017). There were no statistically significant differences in other molecular alterations among favorable and unfavorable 3GES risk cases.

Lastly, we evaluated the tumor microenvironment through computational immune cell deconvolution. Surprisingly, the tumors of patients from the unfavorable 3GES risk category were infiltrated by a significantly higher proportion of immune cells such as B cells (*P* = 0.036), CD8+ T cells (*P* = 0.004), and macrophages (*P* < 0.001) (**Figure 3A**). The estimated proportion of NK cells was negligible for all samples. To explore further this, we examined the level of expression of different immune exhaustion markers on these tumor samples. Compared to tumors from patients of the favorable 3GES risk category, those from the unfavorable-risk group presented a significantly higher expression of immune exhaustion markers such as *CTLA4* (*P* = 0.001), *LAG3* (*P* < 0.001), *PDCD1* (*P* = 0.023), and *TIGIT* (*P* < 0.001) (**Figure 3B**). Consistently with this finding and with the clinical value of our 3GES, those tumors from the unfavorable-risk group exhibit a significantly higher proportion of cancer associated fibroblasts (CAFs) (*P* = 0.003) (**Figure 3B**).

**Figure 3.**
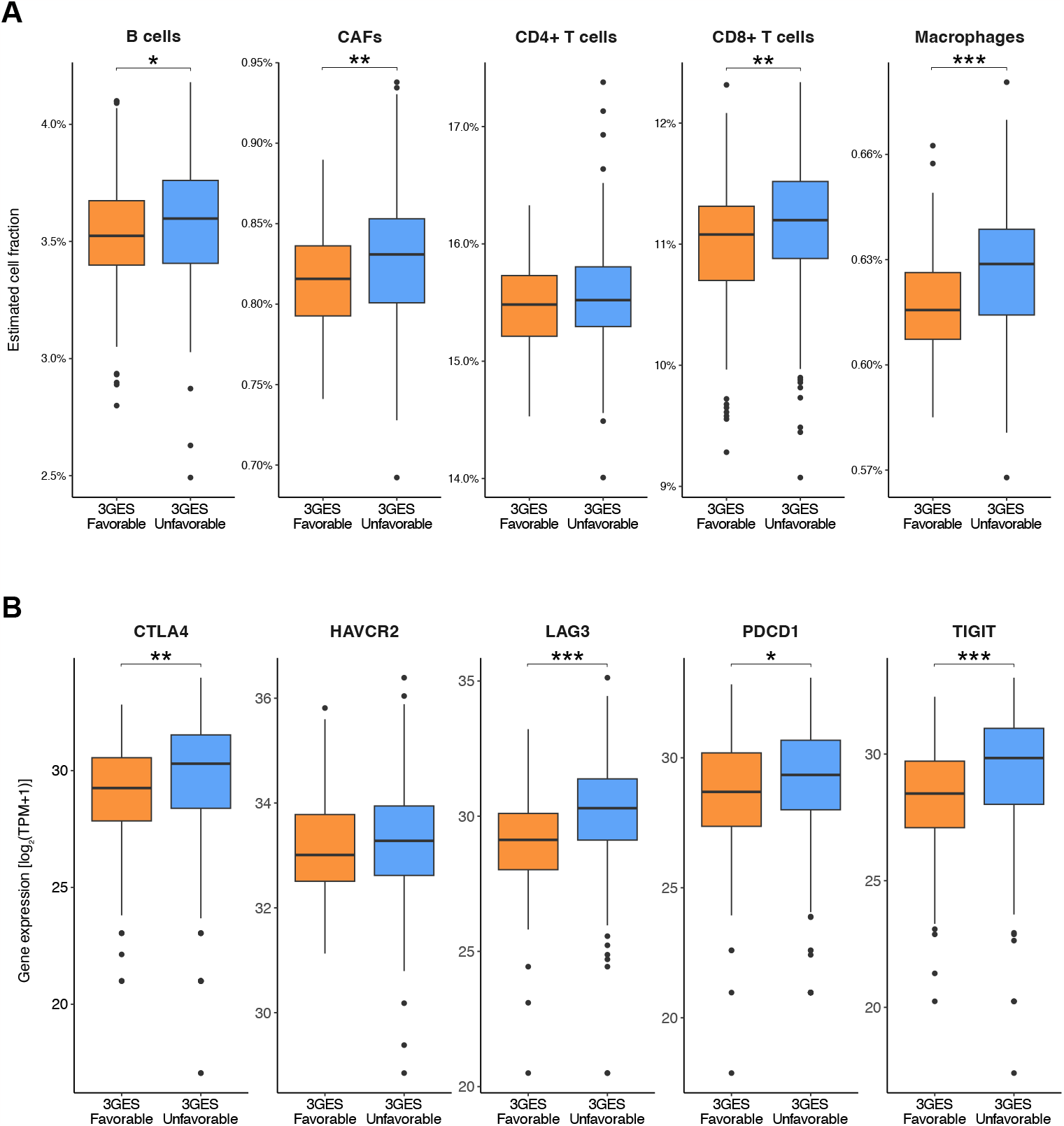
Comparisons between **(A)** estimated cell fractions and **(B)** immune exhaustion markers expression levels by 3GES among patients from the pooled cohort of the CheckMate-009, -010, and - 025 trials. In the boxplots, whiskers represent the variability outside the upper and lower quartiles, the middle line in the box stands for median, the bounds of the box stand for upper and lower quartiles, and dots stand for outliers. *P* values of Wilcoxon rank-sum tests within pairs of gene groups are indicated by asterisks (\**P* < 0.05, \*\**P* < 0.01, \*\*\**P* < 0.001). Abbreviations: 3GES, 3-gene expression score; CAFs, cancer-associated fibroblasts; TPM, transcripts per million.

## Discussion

Nowadays, in the accRCC scenario there are no consistent ICB predictive biomarkers. Even though multiple studies have been conducted to discover predictive biomarkers in this context, only IMDC Risk Score is used in clinical practice to select those patients who are candidates for treatment with the combination regimen nivolumab plus ipilimumab^7^. Moreover, although microsatellite instability and TMB are also FDA-approved as predictive biomarkers for pembrolizumab in a tissue/site-agnostic cancer indication, its utility in ccRCC is anecdotal and debatable. Taking this into consideration, herein we conducted a retrospective study in order to develop and to validate a gene expression score for predicting clinical benefit to the anti-PD-1 antibody nivolumab in the context of patients diagnosed with accRCC enrolled in the CheckMate-009, -010 and -025 clinical trials. Additionally, we explored the correlation of our 3GES with different clinical, molecular, and immune tumor characteristics.

First, we systematically developed a novel 3GES with prognostic value in a pooled cohort of 181 patients treated with nivolumab from the CheckMate-009, -010 and -025 trials. Considering the expression level of 3 independent prognostic genes, we developed a simple model to segregate patients into two risk categories based on the hazard of death (favorable and unfavorable prognostic groups), which importantly was also associated with DC and response. Next, we evaluated and validated the prognostic significance of our score in an independent dataset, the TCGA-KIRC cohort. Regarding the genes included in the risk score, none of them have been previously described as either a prognostic or predictive biomarker in RCC. However, in vitro functional analysis in human RCC cell lines specifically revealed that HMGA1 knockdown markedly inhibited colony formation, significantly induced apoptosis, inhibited invasion potential and induced anoikis, suggesting this molecule as a potential target for novel therapeutic modalities for advanced RCC^25^. No relevant information is available related to the functional role in RCC of the other evaluated transcripts.

Second, once the prognostic value of our 3GES was confirmed, we further evaluated its capacity as a predictive biomarker for anti-PD-1 blockade. For this purpose, we specifically interrogated our score among patients enrolled in the CheckMate-025 study, a two-arm, randomized, open-label, phase 3 study of nivolumab in comparison with everolimus. The 3GES showed a statistically significant interaction with the treatment arm; while among patients with an unfavorable 3GES risk there was not a significant difference in terms of OS based on the allocated treatment arm, among those with a favorable score nivolumab monotherapy significantly improved the OS compared to everolimus.

Lastly, according to clinical and pathological features, we found an enrichment of different characteristics classically associated with better prognosis among those cases with a favorable 3GES risk such as a favorable MSKCC risk score^26^ and the absence of sarcomatoid and/or rhabdoid histological differentiation^27,28^. Moreover, when we evaluated the molecular profile of tumors, we found a higher proportion of PBRM1 loss-of-function mutations and a lower proportion of the amplification 8Q24.3 among those cases with a favorable 3GES risk. Although initially there was evidence supporting the role of *PBRM1* loss-of-function mutations as a positive predictive biomarker for the anti-PD-1 antibody nivolumab^9,22,29^, new data claiming the opposite role^30^ has hampered its translation to the clinic. Interestingly, regarding the amplification of 8Q24.3, Braun et al reported a higher frequency of this molecular alteration among immune-infiltrated ccRCC tumors^9^. Additionally, favorable 3GES risk cases presented a lower wGII, a genomic characteristic previously associated with a less aggressive phenotype compared to those ccRCC tumors with a higher wGII^31^. From an immune perspective, when we evaluated the tumor microenvironment through computational immune cell deconvolution found that those tumors from patients with a favorable 3GES risk overall presented lower immune cell infiltrates compared with their counterparts with an unfavorable 3GES risk. However, despite having higher levels of immune cell infiltration, unfavorable 3GES risk tumors presented a higher expression of immune exhaustion markers, which could explain their worse clinical outcome. Furthermore, patients from the unfavorable 3GES risk group exhibited a higher proportion of CAFs, a fibroblast population with a well-documented immunosuppressive role which limits anti-PD-1 blockade efficacy^32,33^. Based on this, one could hypothesize that patients with unfavorable 3GES risk tumors would benefit from combination strategies against CTLA-4 and other non-classical immune checkpoint molecules such as LAG-3 or TIGIT. Moreover, due to the higher proportion of CAFs, emerging therapies targeting this cellular population by either depleting them, reducing their tumor-promoting and immunosuppressive functions or even by reprogramming them to a more quiescent state^34^, are potential strategies to improve clinical outcomes in this population characterized by an unfavorable 3GES.

Our study has two main limitations. First, it pivots on a *post hoc* pooled analysis of those patients from the clinical trials CheckMate-009, -010 and -025 with enough clinical and molecular tumor data, and who consented to participate. Though Braun et al have reported that these patients do not differ significantly with respect to survival from the whole population enrolled in the trials^9^, this fact could lead to an uncontrolled selection bias. On a positive note, the availability of a control arm of patients treated with everolimus in the CheckMate-025 has allowed us to explore the predictive nature of our 3GES through the evaluation of biomarker-treatment interaction with the likelihood ratio test. The second limitation is the use of optimal cutoff thresholds to define our genes as high or low based on RNA-seq data. While promising, to confirm the utility of 3GES in a daily clinical practice scenario, our results should be validated with an easy to implement orthogonal technique such as RT-qPCR or nCounter assays. Furthermore, measuring immune cells from RNA expression data can be error prone. Therefore, computational immune cell deconvolution results, although consistent, need to be interpreted cautiously in the absence of an independent validation method.

Today, either in daily clinical practice or in a clinical trial scenario, there are available different treatment options for the management of patients with accRCC. In this context, the development of tools to help in the decision-making process is mandatory. In this study, in addition to develop and to validate a 3GES for predicting clinical benefit to the anti-PD-1 antibody nivolumab in the context of patients diagnosed with accRCC, we characterized its underlying clinical, molecular, and immune features. If the results of this study are definitively validated in other retrospective and large-scale, prospective studies, the 3GES will represent a valuable tool for guiding the design of ICB-based clinical trials in this scenario in the near future.

## Data Availability

All clinical, molecular, and immune tumor data from CheckMate-009, -010, and -025 trials used for this retrospective study have been made freely available through Supplementary Material by Braun et al (https://www.nature.com/articles/s41591-020-0839-y).

## Acknowledgements

YZB is supported by a predoctoral fellowship from Xunta de Galicia (ED481A 2022/491). MF-P is supported by a Santander Investigación predoctoral research contract from the University of Santiago de Compostela. MK is supported by a predoctoral fellowship from Xunta de Galicia (ED481A 2022/422). JR-B is supported by a Juan Rodés contract (JR21/00019) from the Institute of Health Carlos III. The results shown here are in part based upon data generated by Braun et al (https://www.nature.com/articles/s41591-020-0839-y) and the TCGA Research Network (https://www.cancer.gov/tcga). We thank the Supercomputing Centre of Galicia (CESGA) for providing complementary computational resources.

## Author Contributions

Conceptualization: YZB, MF-P, and JR-B; software, YZB and JR-B; validation, YZB and JR-B; formal analysis, YZB and JR-B; investigation: all authors; resources: all authors; data curation, all authors; writing – original draft, YZB and JR-B; writing – review & editing, all authors; visualization, YZB, and JR-B; supervision, JR-B; project administration, J.R.-B; funding acquisition, JR-B.

## Conflicts of interest

Urbano Anido-Herranz—Travel, accommodations, expenses: Ipsen, Bayer, Merck, Pfizer, and Sanofi; honoraria for educational activities: Advanced Accelerator Applications - Novartis, Bayer, Ipsen, MSD, AstraZeneca, Merck, Eisai, Bristol-Myers Squibb, Kyowa Kirin, Rovi, GlaxoSmithKline, and LEO Pharma; honoraria for consultancies: Advanced Accelerator Applications - Novartis, Ipsen, AstraZeneca, Merck, Pfizer, Astellas, and Bayer.

Mar Fuentes-Losada—Travel, accommodations, expenses: Seagen.

Victor Cebey-López—Travel, accommodations, expenses: AstraZeneca, Bristol-Myers Squibb, Eisai, Ipsen, Kyowa Kirin, Merck, Novartis, Pfizer, Pharmamar, Pierre-Fabre, Roche, and Sanofi; honoraria for educational activities: AstraZeneca and Pharmamar.

Luis León-Mateos—Travel, accommodations, expenses: Bristol-Myers Squibb, Lilly, MSD, and Roche; honoraria for educational activities: AstraZeneca, Boehringer Ingelheim, Novartis, Jansen, Astellas, and Sanofi; honoraria for consultancies: AstraZeneca, Boehringer Ingelheim, Novartis, Jansen, Astellas, and Sanofi.

Jorge García-González—Travel, accommodations, expenses: AstraZeneca, Bristol-Myers Squibb, MSD, Roche, Sanofi, and Takeda; honoraria for educational activities: AstraZeneca, Bristol-Myers Squibb, MSD, Novartis, Pierre-Fabre, Roche, Sanofi, and Takeda; honoraria for consultancies: AstraZeneca, Boehringer Ingelheim, Bristol-Myers Squibb, MSD, Novartis, Roche, Sanofi, and Takeda.

Natalia Fernández-Díaz—Travel, accommodations, expenses: GlaxoSmithKline, Lilly, Roche, and Sanofi.

Rafael López-López—Travel, accommodations, expenses: Lilly, Novartis, Pfizer, Merck, Roche, and Bristol-Myers Squibb; honoraria for educational activities: Lilly, Novartis, Pfizer, Merck, Roche, and Bristol-Myers Squibb; honoraria for consultancies: Pharmamar, Bayer, and Pierre Fabre.

Juan Ruiz-Bañobre—Travel, accommodations, expenses: Merck, Pierre-Fabre, Sanofi, and Seagen; honoraria for educational activities: Ipsen; institutional research funding: Nouscom, Pfizer, and Roche. Yoel Z. Betancor, Rafael López-López, and Juan Ruiz-Bañobre are inventors in one patent application over these results.

The other authors have no conflicts of interest to declare.

