## Supplementary Material for "A three-gene expression score for predicting clinical benefit to anti-PD-1 blockade in advanced renal cell carcinoma"

8 Co-senior authors.

**[#]Corresponding author:**

### Supplementary Material

**Supplementary Tables:**

- Supplementary Table 1
- Supplementary Table 2
- Supplementary Table 3
- Supplementary Table 4
- Supplementary Table 5

**Supplementary Table 1.** Univariable Cox regression analyses for overall survival considering baseline clinicopathological characteristics among nivolumab treated patients included in the pooled cohort of the CheckMate-009, -010, and -025 trials.

| **Characteristic**[a] | **HR (95% CI)** | ***P*** |
|---|---|---|
| Sex (male vs female) | 1.69 (1.09 - 2.63) | 0.019 |
| Age (> vs ≤ median) | 0.82 (0.57 - 1.18) | 0.288 |
| MSKCC risk (favorable vs unfavorable) | 0.44 (0.29 - 0.68) | < 0.001 |
| Sarcomatoid or rhabdoid (yes vs no) | 1.73 (1.03 - 2.89) | 0.039 |
| Number of prior therapies (continuous) | 1.23 (0.87 - 1.45) | 0.360 |

[a]Baseline variables IMDC risk and site of metastases were excluded from univariable Cox regression analyses because these data were not available in most cases.

Abbreviations: CI, confidence interval; HR, hazard ratio; MSKCC, Memorial Sloan Kettering Cancer Center.

**Supplementary Table 2.** Univariable Cox regression analyses for overall survival among nivolumab treated patients included in the pooled cohort of the CheckMate-009, -010, and -025 trials.

| **Characteristic** | **HR (95% CI)** | **FDR-adjusted *P*** |
|---|---|---|
| *SPOCD1* (high *vs* low) | 3.14 (2.11 - 4.67) | < 0.001 |
| *MYO9B* (high *vs* low) | 4.17 (2.49 - 6.98) | 0.003 |
| *AUP1* (high *vs* low) | 2.72 (1.90 - 3.91) | 0.003 |
| *HMGA1* (high *vs* low) | 2.73 (1.89 - 3.95) | 0.004 |
| *HIST2H2BF* (high *vs* low) | 3.27 (2.11 - 5.07) | 0.006 |
| *USF1* (high *vs* low) | 3.15 (2.04 - 4.85) | 0.009 |
| *NUP62* (high *vs* low) | 2.59 (1.80 - 3.71) | 0.010 |
| *ARHGAP42* (high *vs* low) | 0.39 (0.27 - 0.56) | 0.017 |
| *ADCY5* (high *vs* low) | 0.38 (0.26 - 0.56) | 0.017 |
| *MOCS1* (high *vs* low) | 0.36 (0.25 - 0.54) | 0.022 |
| *RP5_1107A17.2* (high *vs* low) | 2.95 (1.93 - 4.51) | 0.025 |
| *PSMD10P2* (high *vs* low) | 3.56 (2.16 - 5.87) | 0.026 |
| *CLIC4* (high *vs* low)) | 0.30 (0.18 - 0.48) | 0.035 |
| *CDH5* (high *vs* low) | 0.37 (0.25 - 0.55) | 0.039 |
| *HMMR* (high *vs* low) | 2.85 (1.87 - 4.34) | 0.044 |
| *HSF2BP* (high *vs* low) | 2.50 (1.73 - 3.61) | 0.048 |
| *DIAPH3* ((high *vs* low) | 2.75 (1.83 - 4.14) | 0.049 |

Abbreviations: CI, confidence interval; FDR, False Discovery Rate; HR, hazard ratio.

**Supplementary Table 3.** Univariable logistic regression analyses for disease control and response among nivolumab treated patients included in the pooled cohort of the CheckMate-009, -010, and -025 trials.

| | Disease control | | Response | |
|---|---|---|---|---|
| **Characteristic** | **OR (95% CI)** | ***P*** | **OR (95% CI)** | ***P*** |
| *MYO9B* (high *vs* low) | 0.28 (0.10 - 0.77) | 0.014 | 0.37 (0.08 - 1.68) | 0.199 |
| *AUP1* (high *vs* low) | 0.45 (0.25 - 0.84) | 0.011 | 0.57 (0.26 - 1.23) | 0.153 |
| *HMGA1* (high *vs* low) | 0.57 (0.30 - 1.07) | 0.080 | 0.25 (0.09 - 0.69) | 0.007 |
| *USF1* (high *vs* low) | 0.29 (0.13 - 0.66) | 0.003 | 0.21 (0.05 - 0.93) | 0.039 |
| *NUP62* (high *vs* low) | 0.48 (0.26 - 0.89) | 0.019 | 0.53 (0.24 - 1.17) | 0.116 |
| *ARHGAP42* (high *vs* low) | 2.17 (1.15 - 4.09) | 0.016 | 2.17 (0.93 - 5.08) | 0.074 |
| *MOCS1* (high *vs* low) | 2.36 (1.18 - 4.72) | 0.015 | 1.61 (0.66 - 3.96) | 0.299 |

Abbreviations: CI, confidence interval; OR, odds ratio.

**Supplementary Table 4.** Multivariable Cox regression analyses for overall survival among nivolumab treated patients from the pooled cohort of the CheckMate-009, -010, and -025 trials.

| **Characteristic** | **HR (95% CI)** | ***P*** |
|---|---|---|
| *MYO9b* (high *vs* low) | 1.43 (0.72 - 2.88) | 0.310 |
| *AUP1* (high *vs* low) | 1.40 (0.91 - 2.17) | 0.127 |
| *HMGA1* (high *vs* low) | 1.60 (1.05 - 2.46) | 0.031 |
| *USF1* (high *vs* low) | 1.51 (0.85 - 2.65) | 0.157 |
| *NUP62* (high *vs* low) | 1.74 (1.17 - 2.60) | 0.007 |
| *ARHGAP42* (low *vs* high) | 1.74 (1.15 - 2.63) | 0.008 |
| *MOCS1* (high *vs* low) | 1.58 (1.00 - 2.52) | 0.051 |

Abbreviations: CI, confidence interval; HR, hazard ratio.

**Supplementary Table 5.** Baseline patient and disease characteristics of 311 patients included in the pooled cohort of the CheckMate-009, -010, and -025 trials.

| **Characteristic** | **Total** (n = 311) | **Favorable 3GES** (n = 123, 40%) | **Unfavorable 3GES** (n = 188, 60%) | ***P*** |
|---|---|---|---|---|
| **Clinical trial (%)** | | | | 0.913 |
| CM-009 and -010 | 61 | 25 (20) | 36 (19) | |
| CM-025 | 250 | 98 (80) | 152 (81) | |
| **Age (%)** | | | | 0.051 |
| Age > 63 years (median) | 141 | 65 (53) | 76 (41) | |
| Age ≤ 63 years | 168 | 58 (47) | 110 (59) | |
| NA | 2 | | | |
| **Sex (%)** | | | | 0.778 |
| Female | 82 | 34 (28) | 48 (26) | |
| Male | 229 | 89 (72) | 140 (74) | |
| **MSKCC risk (%)** | | | | 0.039 |
| Favorable | 100 | 48 (41) | 52 (29) | |
| Unfavorable | 195 | 68 (59) | 127 (71) | |
| NA | 16 | | | |
| **Sarcomatoid and/or rhabdoid differentiation (%)** | | | | 0.042 |
| No | 247 | 103 (92) | 144 (83) | |
| Yes | 39 | 9 (8) | 30 (17) | |
| NA | 25 | | | |
| **Ploidy (%)** | | | | 1 |
| Ploidy > 1.98 (median) | 91 | 37 (46) | 54 (45) | |
| Ploidy ≤ 1.98 | 108 | 43 (54) | 65 (54) | |
| NA | 112 | | | |
| **Immunophenotype (%)** | | | | 0.290 |
| Desert or excluded | 24 | 13 (30) | 11 (19) | |
| Infiltrated | 79 | 31 (70) | 48 (81) | |
| NA | 208 | | | |
| **wGII (%)** | | | | 0.035 |
| wGII > 0.11 (median) | 99 | 32 (40) | 67 (56) | |
| wGII ≤ 0.11 | 100 | 48 (60) | 52 (44) | |
| NA | 112 | | | |
| **ITH (%)** | | | | 0.053 |
| ITH > 0.26 (median) | 99 | 47 (59) | 52 (44) | |
| ITH ≤ 0.26 | 100 | 33 (41) | 67 (56) | |
| NA | 112 | | | |

*P* values were computed excluding NA cases.

Abbreviations: CM, CheckMate; MSKCC, Memorial Sloan Kettering Cancer Center; ITH, Intra-Tumor Heterogeneity; NA, not available; wGII, weighted Genome Instability Index.
